# Sex-specific Risk Factors for Survival in B-cell Non-Hodgkin Lymphoma Patients after Anti-CD19 CAR T-Cell Therapy

**DOI:** 10.1101/2024.12.12.24318935

**Authors:** Manishkumar S. Patel, Agrima Mian, Akansha Jalota, Peter Bazeley, Sujata Patil, Brian T. Hill, Neetu Gupta

**Affiliations:** Department of Inflammation and Immunity, Lerner Research Institute; Department of Hematology and Medical Oncology, Taussig Cancer Institute; Department of Quantitative Health Sciences, Lerner Research Institute; Department of Cancer Biostatistics, Taussig Cancer Institute, Cleveland Clinic, Cleveland, OH

**Keywords:** CAR T-cells, lymphoma, survival, biological sex

## Abstract

Sex bias is well documented in autoimmune diseases, cancer and immune responses to infectious agents. Here, we investigated if pre-treatment risk factors that influence the survival of B-cell non-Hodgkin lymphoma (NHL) patients after anti-CD19 CAR T-cell therapy are sexually dimorphic. We measured pre-leukapheresis tumor burden (lactate dehydrogenase levels), C-reactive protein (CRP) and serum cytokine and chemokine concentration in 67 B-cell NHL patients treated with axicabtagene ciloleucel (axi-cel) or tisagenlecleucel (tisa-cel). Association of relative abundance of each factor with progression-free survival (PFS) and overall survival (OS) was analyzed in male and female patients together, or only within the male cohort or only within the female cohort. No differences in PFS or OS or in pre-treatment tumor burden, CRP and cytokine/chemokine levels were observed between male and female patients undergoing axi-cel or tisa-cel therapy. However, within the male group, patients with higher pre-treatment tumor burden and greater relative abundance of CRP and pro-inflammatory cytokines and chemokines conferred greater risk of poor progression-free survival (PFS) and/or overall survival (OS). In contrast, within the female group, patient survival was largely agnostic to variations in tumor burden, CRP and cytokine/chemokine abundance. Specifically, higher relative abundance of IL-6, IL-8, IL-27, TNF-α, Eotaxin-1, MIP-1β and MCP-1 was associated with poor PFS and/or OS after CAR T-cell therapy within the male group, whereas higher IL-27 and IFNα2 abundance was associated with better PFS and poorer OS, respectively, within the female group. Our data suggest that biological sex may modulate the impact of baseline risk factors on survival outcomes of CAR T-cell therapy in B-cell NHL.

## Introduction

Anti-CD19 chimeric antigen receptor (CAR) T-cell therapy demonstrates remarkable efficacy for the treatment of refractory/relapsed (r/r) large B-cell lymphoma (LBCL), follicular lymphoma and mantle cell lymphoma^1–3^. However, ∼40-60% of patients treated with CAR-T cell therapy exhibit poor response and survival^1–4^. This has spurred the need to identify pre-treatment biomarkers of response to anti-CD19 CAR T-cell therapy. In addition to tumor characteristics and intrinsic composition/attributes of CAR-T cells, the inter-patient heterogeneity in immunological signatures may influence response to this therapy. Further, factors that do not directly stem from the immune system, such as sex, race and ethnicity may play a role in modulating and/or predicting the survival of patients undergoing CAR T-cell therapy. Indeed, race and ethnicity were recently shown to contribute to the differences in response to CAR T-cell therapy^5^. However, as race and ethnicity are more complex to define, self-reported race is widely recognized as an incomplete interpretation of genetic ancestry or human genetic variation^6^. Further, as reporting these variables is not mandatory^7^ and complete medical records for race and ethnicity are not available for all patients seen at Cleveland Clinic, the present study is unable to address the impact of race and ethnicity on outcomes.

On the other hand, it is well-established that the male and female immune systems are different, and that immune responses to infections^8^, many autoimmune diseases^9–11^, and tumor pathology^12,13^ exhibit sex bias. Sex differences can alter the tumor microenvironment^14^ and modulate immune system homeostasis and responses via inflammatory cytokine production^15, 16^. The impact of sex has also been reported in the efficacy^17^ and survival outcomes of patients^18^ receiving immune checkpoint inhibitors for solid tumors. Therefore, baseline tumor burden and cytokine/chemokine profiles in male and female patients may differentially impact the survival outcomes of anti-CD19 CAR T-cell therapy. Understanding these differences may provide insights into sex-specific biomarkers of survival and enable better stratification of outcomes. As biological sex at birth is reported for all patients undergoing anti-CD19 CAR T-cell therapy, we focused on investigating potential differences in outcomes based on baseline features of responding and non-responding male and female cohorts.

We examined if baseline cytokine/chemokine levels and tumor burden affect the survival of LBCL patients treated with *axicabtagene ciloleucel* (axi-cel) or *tisagenlecleucel* (tisa-cel) in a sex-specific manner. We demonstrate that male B-NHL patients with elevated baseline abundance of IL-8, IL-6, IL-27, Eotaxin-1, MIP-1β, MCP-1, TNF-α and CRP, as well as higher pre-treatment tumor burden exhibit poorer survival compared to those with lower abundance of these factors. On the other hand, within the female patient cohort, the survival outcomes are largely unaffected by the relative abundance of these factors. Our data suggest that an unfavorable baseline circulating cytokine/chemokine profile and high pre-treatment tumor burden may collaborate to lower the survival of male patients undergoing anti-CD19 CAR T-cell therapy.

## Materials and Methods

### Patient enrollment, specimen collection protocol, and clinical data

All patients with r/r B-cell NHL (n=67) who received treatment with axi-cel (n=45) or tisa-cel (n=22) between 2018 and 2021 and gave informed consent for blood sample collection were included in this IRB-approved study, which was executed in accordance with the declaration of Helsinki. In addition to routine blood collection for clinical care, peripheral blood was also collected on the day of leukapheresis (baseline) and serum was stored for inclusion in the institutional biorepository. The patient cohort is represented by 46 males and 21 females and represents the observed male/female ratio within the B-NHL population^19,20^. Baseline clinical characteristics and demographic details were retrieved through an IRB-approved retrospective chart review of the electronic medical record system and maintained in a secure and HIPPA-compliant Research Electronic Data Capture (REDCap) system. Clinical outcomes including progression-free survival (PFS) and overall survival (OS) were recorded relative to the date of CAR T-cell infusion. Patients who did not experience the event by the end of the study period or lost to follow-up were censored for these time-to-event outcomes. Circulating lactate dehydrogenase (LDH) and C-reactive protein (CRP) levels were measured on the day of leukapheresis.

### Cytokine/Chemokine and tumor burden quantification

Multiplexed bead arrays (Eve Technologies, Alberta, Canada) were used to quantify serum levels of 29 cytokines and chemokines, including IL-1RA, IL-3, IL-4, IL-5, IL-6, IL-7, IL-8, IL-9, IL-10, IL-15, IL-18, IL-21, IL-27, IFNα2, IFN-γ, TNF-α, Eotaxin-1, MIP-1α, MIP-1β, MCP-1, Flt3L, Fractalkine, G-CSF, GM-CSF, IP-10, MDC, RANTES, sCD40L and VEGF-A.

### Data processing and quality control

Cytokine/chemokine levels were measured in three batches. Cytokines with >20% of missing values (Out of range-OOR) across patients in each batch were removed. Cytokines with ≤20% missing values were imputed as follows^21^. OOR values were replaced with median values; OOR< (out of range at the lower end) and OOR> (out of range at the higher end) were imputed with half of the minimum observed value across all samples and double of the maximum value observed across all samples, respectively. The concentrations were log-transformed and a PCA plot was generated (Supplemental Fig. 1A) to identify batch effects, using the PCA function in the FactoMineR package. Batch correction was performed using “scale and center” in R and the removal of batch effects was again confirmed by generating a PCA plot (Supplemental Fig. 1B). Batch-corrected relative abundance of each cytokine/chemokine was used for the analysis of association with survival outcomes.

### Assessment of clinical response

Response assessment was performed 3 months after CAR T-cell therapy using the Lugano classification system and defined as a complete metabolic response, partial response, stable disease, or progressive disease, based on FDG uptake on PET-CT^22^. Response groups were defined based on ORR = overall response rate (includes complete response [CR] and partial response (PR)), and NR = no response (includes progressive disease [PD], stable disease [SD], death).

### Association and statistical analysis

These analyses had two main goals: to evaluate differences in tumor burden (i.e. LDH), CRP, and each cytokine/chemokine by sex and to evaluate the association of tumor burden, CRP, and each cytokine/chemokine with OS and PFS by sex. Due to skewness, all cytokine, chemokine, LDH and CRP values were log-transformed prior to analysis. These variables were also made binary at established cutoffs or if no established cutoffs existed, at the sex-specific median. Specifically, for LDH, the indication for tumor burden was calculated using two established cutoffs: one at ≥250 U/l^23^ and the other at ≥400 U/l^24^. For all other cytokines and chemokines, cutoffs at sex-specific medians were used. Differences in each continuous variable by sex was assessed using a two-sample Wilcoxon test. For males and females separately, the association of each binary variable with OS and PFS was assessed using the Kaplan-Meier method and the log-rank test. We note that the number of males vs females in the present cohort differs, and thus comparing significance by sex is not valid. We also note that our analyses and results are hypothesis generating and as such, the findings will require validation in a larger, separate dataset. No adjustments for multiple comparisons are made. All statistical analyses were performed using R v4.1.0 and p-values <0.05 were considered significant.

## Results

Baseline and post-treatment clinical characteristics of the r/r B-NHL patient cohort treated with anti-CD19 CAR T-cells are shown in Table 1. The median age was 63 years (range 25-77), and 68.7% of patients in the cohort were male. The median age of males and females was 64.5 years (range 25-77) and 61 years (range 30-74), respectively. The clinical diagnosis was diffuse large B-cell lymphoma (DLBCL) (n = 46, males=35, females=11), transformed follicular lymphoma (n = 12, males=5, females=7), or primary mediastinal B-cell lymphoma (n = 7, males=5, females=2), and 40.3% of the patients had received prior autologous stem cell transplantation. Axi-cel was received by 45 patients (males=29, females=16) and tisa-cel was received by 22 patients (males=17, females=5). The median durations for overall survival (OS) and progression-free survival (PFS) were 332 days and 134 days, respectively.

**Table 1:**
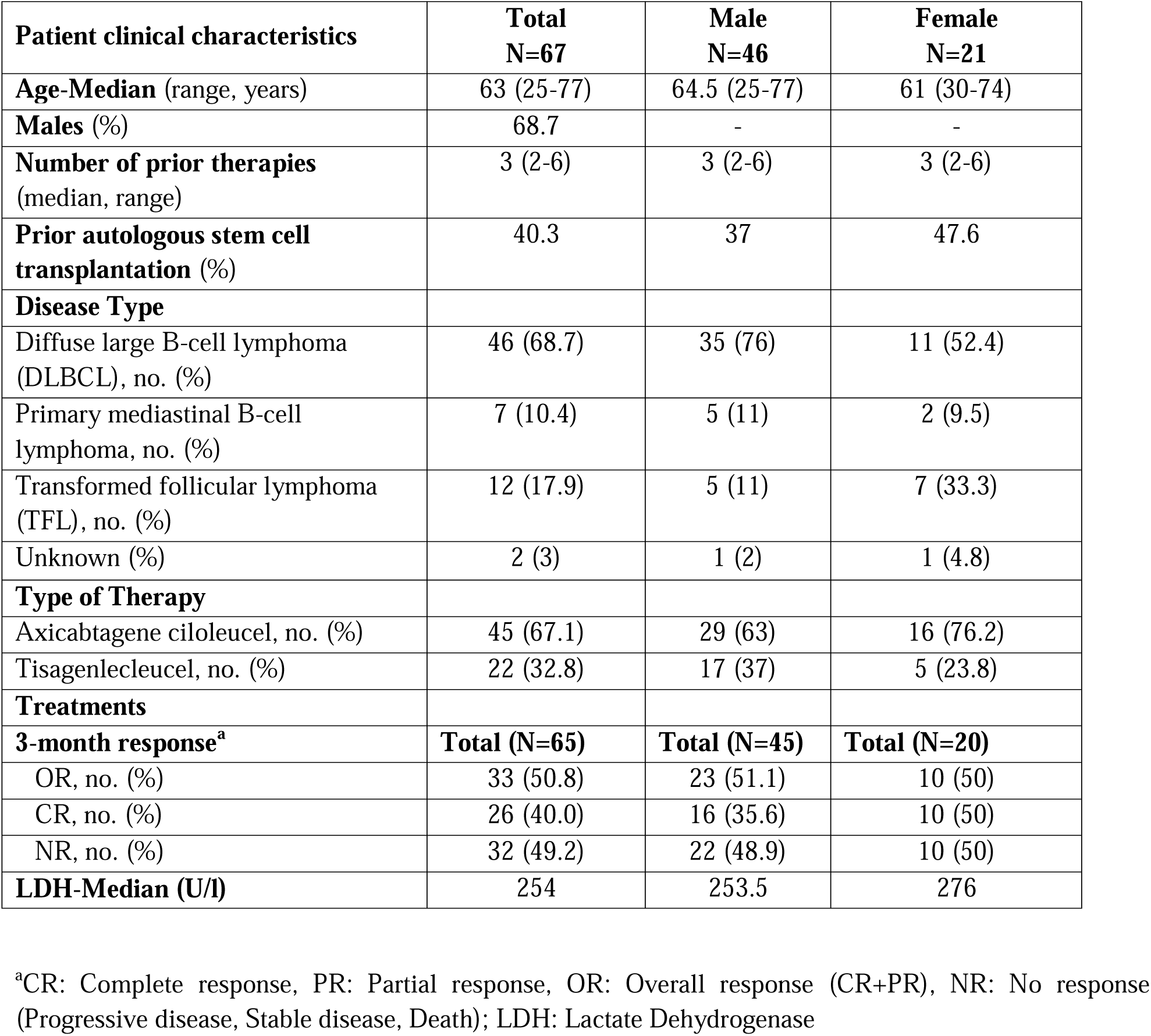
Baseline and post-treatment clinical characteristics.

| **Patient clinical characteristics** | **Total**<br>**N=67** | **Male**<br>**N=46** | **Female**<br>**N=21** |
| --- | --- | --- | --- |
| **Age-Median** (range, years) | 63 (25-77) | 64.5 (25-77) | 61 (30-74) |
| **Males (%)** | 68.7 | - | - |
| **Number of prior therapies**<br>(median, range) | 3 (2-6) | 3 (2-6) | 3 (2-6) |
| **Prior autologous stem cell**<br>**transplantation (%)** | 40.3 | 37 | 47.6 |
| **Disease Type** |  |  |  |
| Diffuse large B-cell lymphoma<br>(DLBCL), no. (%) | 46 (68.7) | 35 (76) | 11 (52.4) |
| Primary mediastinal B-cell<br>lymphoma, no. (%) | 7 (10.4) | 5 (11) | 2 (9.5) |
| Transformed follicular lymphoma<br>(TFL), no. (%) | 12 (17.9) | 5 (11) | 7 (33.3) |
| Unknown (%) | 2 (3) | 1 (2) | 1 (4.8) |
| **Type of Therapy** |  |  |  |
| Axicabtagene ciloleucel, no. (%) | 45 (67.1) | 29 (63) | 16 (76.2) |
| Tisagenlecleucel, no. (%) | 22 (32.8) | 17 (37) | 5 (23.8) |
| **Treatments** |  |  |  |
| **3-month response<sup>a</sup>** | **Total (N=65)** | **Total (N=45)** | **Total (N=20)** |
| OR, no. (%) | 33 (50.8) | 23 (51.1) | 10 (50) |
| CR, no. (%) | 26 (40.0) | 16 (35.6) | 10 (50) |
| NR, no. (%) | 32 (49.2) | 22 (48.9) | 10 (50) |
| **LDH-Median (U/l)** | 254 | 253.5 | 276 |

<sup>a</sup>CR: Complete response, PR: Partial response, OR: Overall response (CR+PR), NR: No response (Progressive disease, Stable disease, Death); LDH: Lactate Dehydrogenase

### High baseline tumor burden and CRP are associated with poor survival in male patients

We first investigated if there is sex bias in overall and progression-free survival of B-cell NHL patients treated with anti-CD19 CAR T cells. Similar to previous findings in pediatric B-ALL patients treated with CD19-directed CAR T-cells^25^, male and female r/r B-cell NHL patients did not show a significant difference in the probability of OS (Supplemental Fig. 2A) or PFS (Supplemental Fig. 2B) upon CAR T-cell therapy. Additionally, the abundance of established baseline risk factors such as tumor burden (measured as LDH) (Supplemental Fig. 2C) and CRP (Supplemental Fig. 2D) were not different between male and female patients in our cohort. Next, we examined if tumor burden and CRP have different associations with survival when the male and female groups and analyzed separately. A previous study used ≥250 U/l LDH to stratify high and low tumor burden and identified this marker as the sole predictor of survival^23^; therefore, initially we first used the same cut-off in our analysis. Patients who had ≥250 U/l LDH prior to treatment were at greater risk of poor PFS and OS after axi-cel (n=45) and tisa-cel (n=22) therapy compared to those with <250 U/l LDH (Fig. 1A). Importantly, while male patients with ≥250 U/l LDH had poorer PFS and OS compared to those with <250 U/l LDH (Fig. 1B), there was no association between LDH concentration and OS or PFS in female patients (Fig. 1C). Next, because ≥400 U/l LDH was reported to be associated with poor PFS in patients receiving anti-CD19 CAR T-cell therapy^24^, we also performed association analysis using this cut-off. We found that ≥400 U/l LDH was significantly associated with poor PFS and OS in all patients, and in the male only group, but not in the female only group (Supplemental Fig. 3). Among pre-treatment circulating biomarkers, CRP is an important regulator of inflammation and elevated serum CRP is known to be associated with poor PFS or OS after CAR T-cell therapy in multiple myeloma patients^26^, and DLBCL patients after CAR T-cell therapy^27^ or chemotherapy^28^. Hence, we further investigated if CRP levels were associated with differential survival outcomes in separate male and female cohorts. Elevated CRP was significantly associated with poor OS when considering both male and female patients together (Fig. 2A, P=0.0172) or males alone (Fig. 2B, P=0.007), but no association was observed in the female only cohort (Fig. 2C, P=0.7336). On the other hand, elevated CRP was not significantly associated with poor PFS in male and female patients together (Fig. 2A, P=0.0932) or only males (Fig. 2B, P=0.1028) or only females (Fig. 2C, P=0.5199). It is important to note that while above median CRP abundance showed a trend of association with poor PFS in male and female patients together (Fig. 2A) and only in males (Fig. 2B), this trend was not evident in the female only cohort (Fig. 2C).

**Figure 1.**
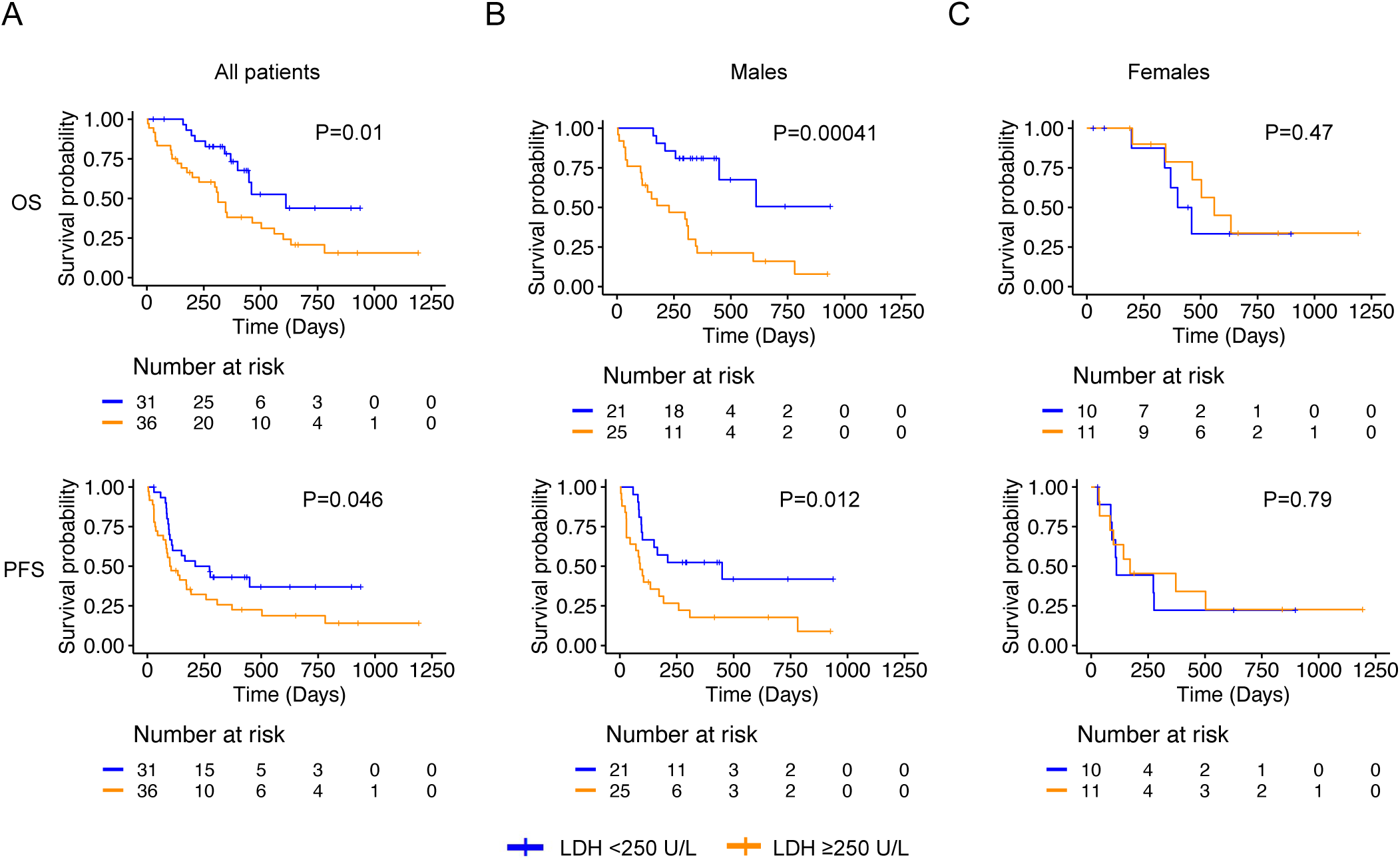
Male patients with higher pre-treatment tumor burden show poorer survival after treated with anti-CD19 CAR T-cells. Association of baseline LDH levels with overall survival (OS), and progression-free survival (PFS) in both male and female **(A)**, only male **(B)**, or only female **(C),** LBCL patients. P<0.05; Males, n=46; Females, n=21; Yes, n=45; Kym, n=22. LDH cut-off, 250 U/l

**Figure 2.**
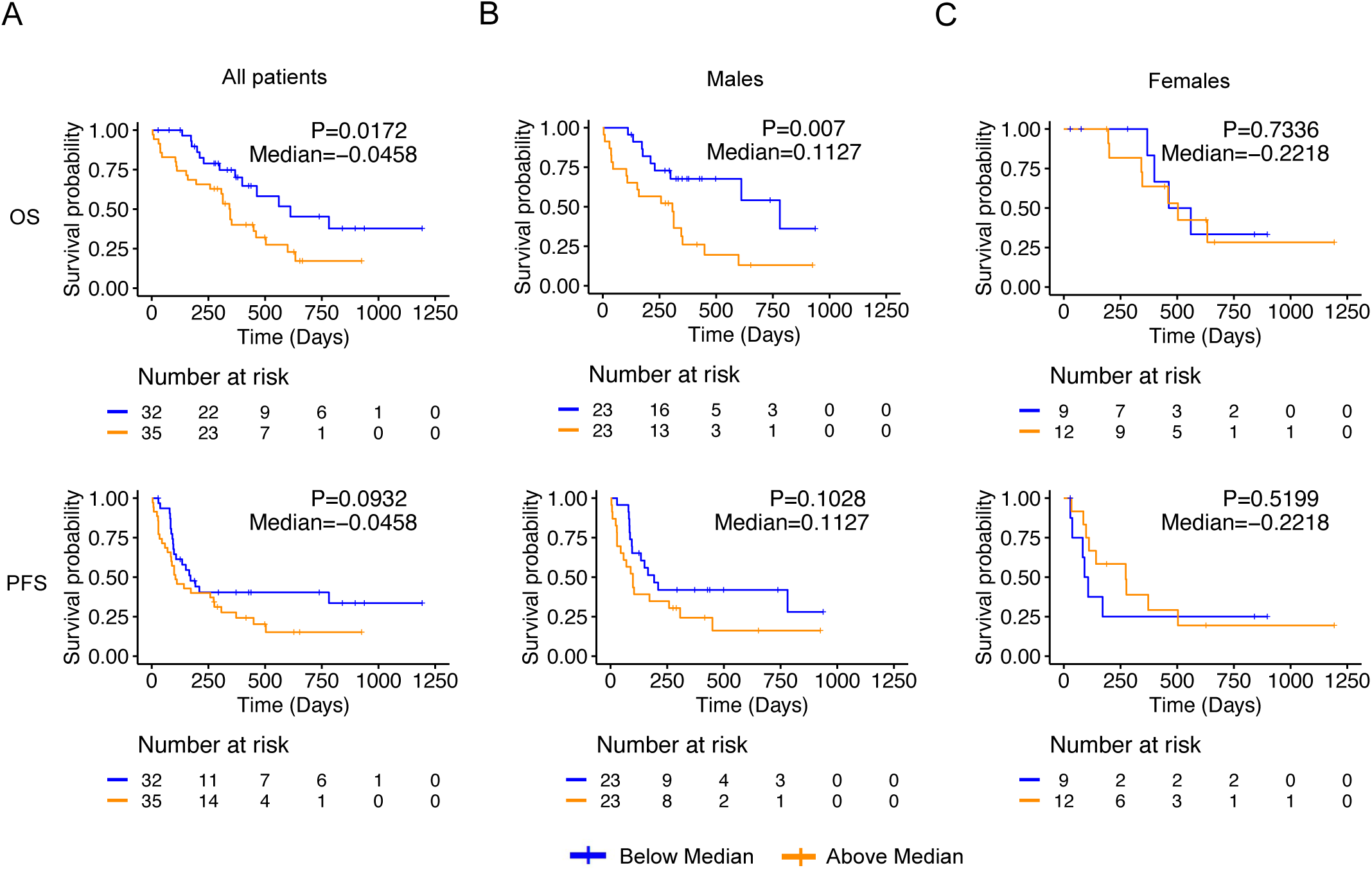
Male patients with higher pre-treatment CRP abundance show poorer survival upon treatment with anti-CD19 CAR T-cells. Association of baseline CRP with overall survival (OS), and progression-free survival (PFS) in both male and female **(A)**, or only male **(B)**, or only female **(C),** LBCL patients. P<0.05; Males, n=46; Females, n=21; Yes, n=45; Kym, n=22.

### High abundance of IL-6, IL-8, IL-27, Eotaxin-1, MIP-1**β** and MCP-1 associated with poor survival in male patients

Because CRP is an important inflammatory biomarker, we examined if the pre-treatment levels of other serum cytokines/chemokines that mediate inflammation during CAR T-cell therapy were different between male and female patients. For this study, we chose 29 cytokines/chemokines (IL-1RA, IL-3, IL-4, IL-5, IL-6, IL-7, IL-8, IL-9, IL-10, IL-15, IL-18, IL-21, IL-27, IFNα2, IFN-γ, TNF-α, Eotaxin-1, MIP-1α, MIP-1β, MCP-1, Flt3L, Fractalkine, G-CSF, GM-CSF, IP-10, MDC, RANTES, sCD40L and VEGF-A) that are of myeloid and lymphoid origin and play essential roles in innate and/or adaptive immunity. With the exception of IL-3, the relative abundance of baseline circulating cytokines and chemokines was not significantly different between male and female patients (Fig. 3). We reasoned that despite no observable differences in abundance, these cytokine/chemokines may have different interactions with the male vs. female immune environment and could therefore be associated differentially with survival in each sex. To investigate this, the association of cytokine abundance with survival was analyzed separately for only male, only female or both sexes together. The relative abundance of specific cytokines/chemokines within each cohort was classified as either below or above median, and median abundance is indicated for each cohort and cytokine in PFS and OS association analysis. When both sexes were analyzed together, poorer PFS was found to be associated with higher abundance of MIP-1β and Eotaxin-1 (P<0.05), and IL-8 (P=0.074) (Fig. 4). When associations of cytokine abundance with PFS were analyzed separately in male only and female only patients, higher abundance of IL-8, Eotaxin-1, MIP-1β (P<0.05), IL-6 (P=0.075), and MCP-1 (P=0.086) were associated with poor PFS in male patients (Fig. 4). The abundance of these cytokines had no observable associations with PFS in the female only cohort but elevated IL-27 was significantly associated with better PFS in females (Fig. 4). Higher relative abundance of IL-6, IL-8, IL-27, Eotaxin-1, MCP-1, and TNF-α was also significantly associated (P<0.05) with poorer OS when considering the male only cohort; but no observable difference in OS was found in the female only cohort for these cytokines. (Fig. 5). Interestingly, IFNα2 was the only cytokine whose higher relative abundance was associated with poor OS (P=0.07) within the female only cohort (Fig. 5). Higher abundance of IL-6 was associated (P<0.05) with poor OS when male and female sexes were analyzed together or in the male only cohort, but no association was observed in the female only cohort (Fig. 5). While IL-7 is an important T-cell promoting cytokine, its pre-treatment abundance was not associated with PFS (Fig. 4) or OS (Fig. 5). Additionally, the relative abundance of IL-1RA, IL-3, IL-4, IL-9, IL-10, IL-15, IL-18, IL-21, Flt3L, Fractalkine, G-CSF, GM-CSF, IP-10, MDC, RANTES, sCD40L, and VEGF-A showed no association with PFS (Supplemental Fig. 4) or OS (Supplemental Fig. 5) in male or female patients. These data indicate that despite similar overall abundance of the analyzed cytokines in male and female r/r B-NHL patients, the association of their individual relative abundance with survival after anti-CD19 CAR T-cell therapy varies in a sex-biased manner. Taken together, our results demonstrate that higher tumor burden, CRP, IL-6, IL-27, TNF-α, IL-8, Eotaxin-1, MIP-1β and MCP-1 represent a novel signature of poor survival in male, but not female B-NHL patients treated with anti-CD19 CAR T-cells.

**Figure 3.**
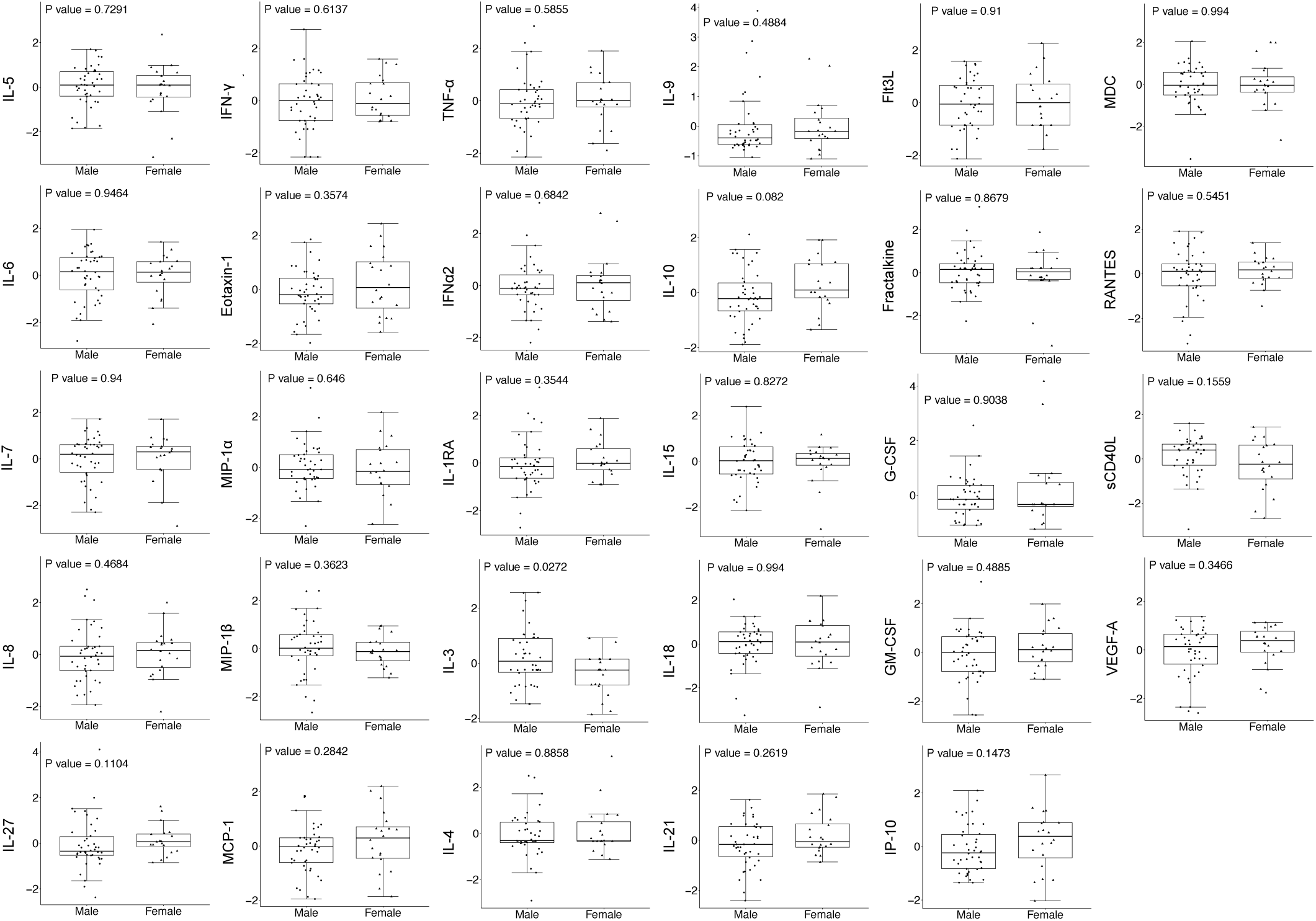
Baseline cytokine and chemokine levels in male and female LBCL patients. Pre-treatment relative abundance of cytokines and chemokines was compared in the serum of male and female LBCL patients. P<0.05; Males, n=42; Females, n=20; Yes, n=42; Kym, n=20.

**Figure 4.**
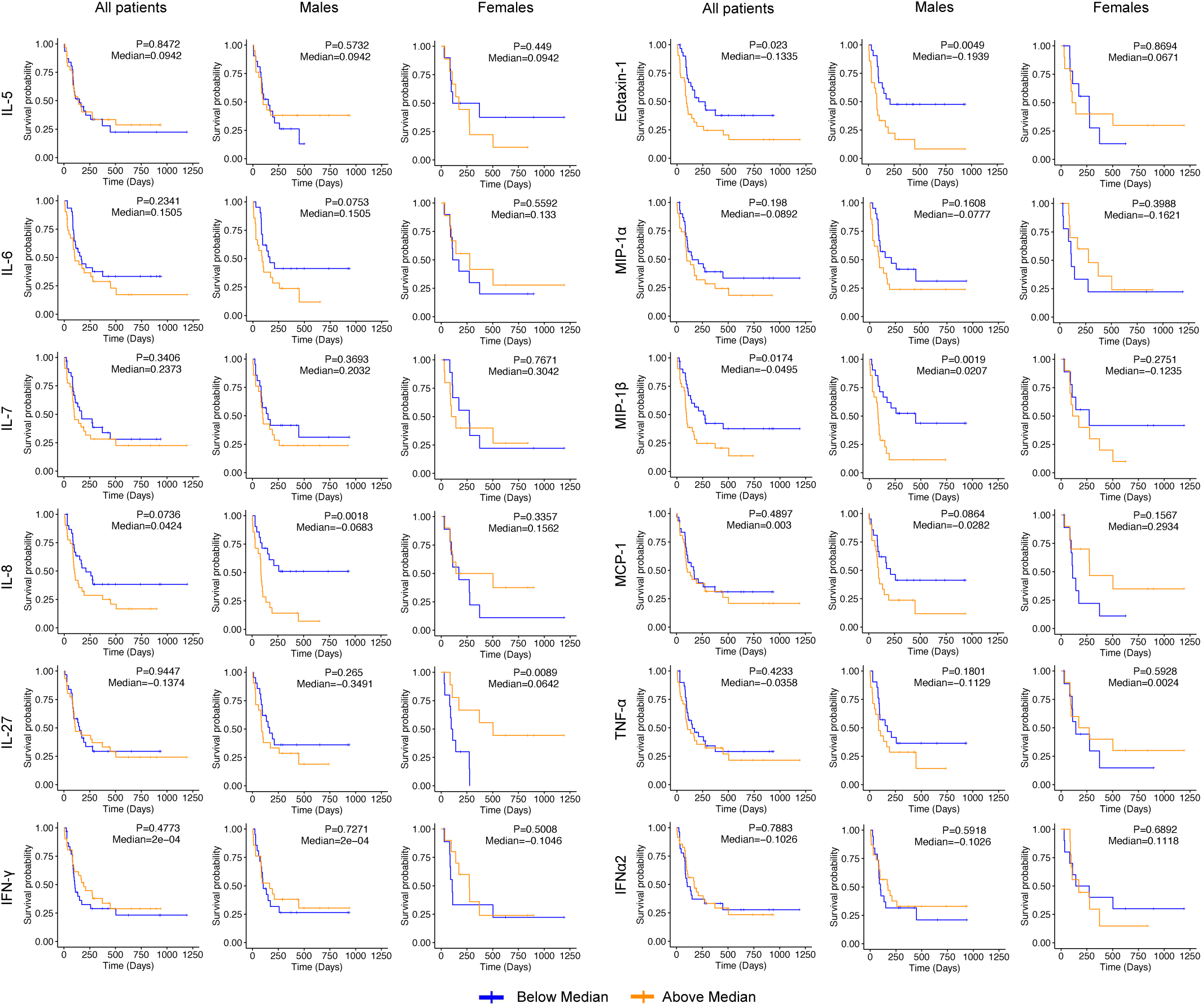
Male patients with elevated pre-treatment abundance of IL-8, Eotaxin-1 and MIP-1β show poorer PFS upon treatment with anti-CD19 CAR T-cells. Association between relative abundance of the indicated cytokines and PFS in both male and female, or only male or only female LBCL patients. P<0.05; Males, n=42; Females, n=20; Yes, n=42; Kym, n=20.

**Figure 5.**
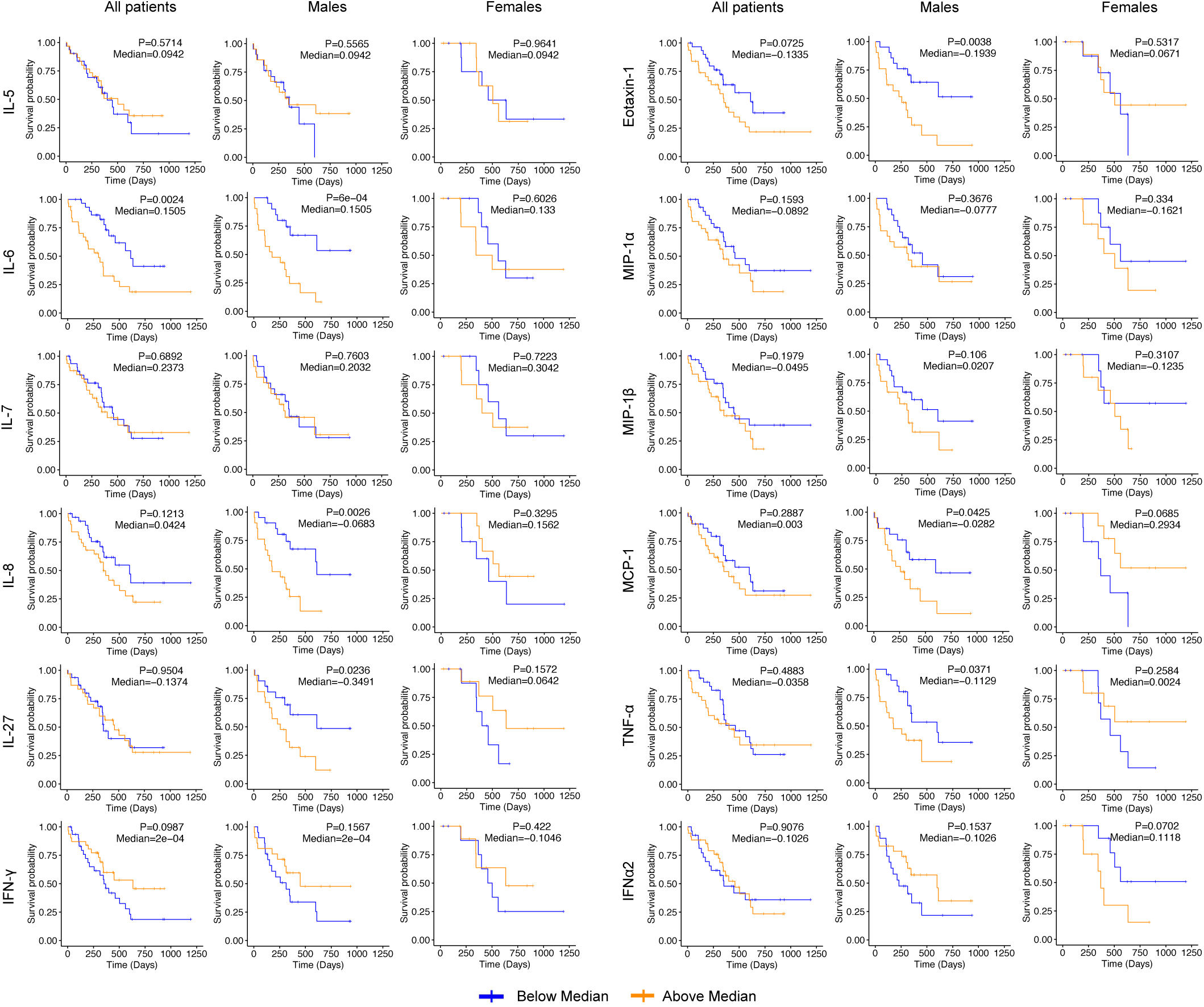
Male patients with elevated pre-treatment abundance of IL-6, IL-8, IL-27, Eotaxin-1, MCP-1 and TNF-α show poorer OS upon treatment with anti-CD19 CAR T-cells. Association between relative abundance of the indicated cytokines and OS in both male and female, or only male or only female LBCL patients. P<0.05; Males, n=42; Females, n=20; Yes, n=42; Kym, n=20.

## Discussion

A variety of pre-treatment biomarkers of clinical outcomes have been reported in LBCL patients undergoing anti-CD19 CAR T-cell therapy^29,30^. However, the impact of sex on the association between pre-treatment tumor and immune environment and survival after CAR T-cell therapy has not been comprehensively explored. Here, we demonstrate that higher baseline tumor burden and greater relative abundance of a pre-treatment inflammatory signature consisting of IL-8, IL-6, TNF-α, Eotaxin-1, MIP-1β, MCP-1 and CRP confers risk of poor survival in male patients treated with anti-CD19 CAR T-cells. On the other hand, female patients undergoing CAR T-cell therapy do not show an association between the relative abundance of these pre-treatment biomarkers and survival. Our findings are consistent with the tenet that the immune system has different interactions in the male and female sex and that this plays a role in establishing sex differences in response to immunotherapy.

A pan-cancer analysis demonstrated sex as a significant factor influencing cancer-specific survival and females have better prognosis than male patients in most cancers^31^. In B-cell lymphoma patients, the male sex is an adverse prognostic factor when treated with immunochemotherapy but not when treated with chemotherapy^32^. Our finding that higher baseline tumor burden predisposes only male patients to a poor survival outcome indicates sex bias in outcomes of CAR T-cell therapy. Specific components of tumors in males may play a role in this predisposition, such as high circulating cytokine levels and monocytic-myeloid-derived suppressor cells (mMDSCs), both of which are associated with resistance to CAR T-cell therapy^30^. Interestingly, males have higher levels of mMDSCs in general^33^ and this is thought to be one of the sex-specific mechanisms that prevents autoimmune diseases and underlies poorer response to infections^34^. Along these lines, CRP, another known biomarker of poor prognosis in r/r DLBCL after CAR T-cell therapy^27^, was reported to support the expansion of MDSCs and suppression of T cell proliferation^35^. It is conceivable that male patients with larger tumor burden have higher levels of mMDSCs which interfere with CAR T-cell activity. Therefore, the association of higher pre-treatment CRP and tumor burden with poor survival suggests the existence of an unfavorable immune environment in male r/r B-NHL patients that may limit optimal CAR T-cell activity.

Inflammatory cytokines and chemokines are important for the activation of T cell-mediated immune reactions and recruitment of myeloid cells, and together these soluble factors facilitate CAR T-cell activity^36–38^. Remarkably, we did not observe a significant difference in the serum abundance of cytokines/chemokines between male and female r/r B-NHL patients treated with anti-CD19 CAR T-cells, with the exception of IL-3. Yet, in male patients, higher pre-treatment abundance of IL-6, IL-8, IL-27, TNF-α, Eotaxin-1 and MIP-1β confers risk of poor survival outcomes. As these associations are not observed in female patients, our findings indicate that the manner in which these cytokines interact with the male environment may be different from that in the female environment, and likely confers poor survival probability in the former. Interestingly, IL-6, TNF-α, IL-8, MIP-1β and MCP-1 are known to be associated with tumor burden and tumor growth in various cancers^39–42^. High tumor burden is associated with the IL-6 signaling pathway^43^ and with elevated levels of TNF-α^44^ and CRP^45^ in aggressive cancers. Additionally, inflammatory stimuli such as IL-6 can indirectly induce the expression of chemokine mediators such as MCP-1^46–48^ which recruit MDSCs and support tumor growth^49^. Eotaxin-1 is a CC subfamily chemokine similar to MCP-1^47^ and is reported to inhibit antitumor immunity^50,51^. Therefore, the association of higher relative abundance of IL-8, IL-6, TNF-α, Eotaxin-1, MIP-1β and MCP-1with poor survival probability in the male patients suggests a unique interaction network between the tumor and circulating environments that predisposes male patients to poorer survival outcomes after anti-CD19 CAR T-cell therapy. Indeed, some of these interactions may be mediated by sex hormones^52^, social behaviors^53,54^, chromosomal genetics^55^, and sex chromosome-encoded genetic environment in the male sex^56^.

While a majority of associations described here impact survival outcomes in male r/r B-NHL patients, there were notable associations between abundance of two cytokines with survival in female r/r B-NHL patients – IL-27 and IFNα2. Female patients with higher relative abundance of IL-27 showed significant association with better PFS, and those with higher abundance of IFNα2 showed poorer OS. IL-27 is a pleiotropic cytokine with both pro- and anti-inflammatory activities and IL-27 expressing T cells demonstrate sustained anti-tumor immunity and cytotoxicity^57^. Interestingly, our data reveal context-specific associations of this cytokine – in male patients elevated abundance increases risk of poor overall survival, yet in female patients, higher abundance of IL-27 predicts better progression-free survival. The molecular basis of this difference is unclear and deserves future investigation. Type I IFNs, specifically IFNα, are a primary source of pathogenesis in autoimmune diseases such as systemic lupus erythematosus that exhibit a strong female sex bias^58^. Therefore, female r/r B-NHL patients who have higher abundance of IFNα2 may experience similar inflammatory pathologies that affect their overall survival after CAR T-cell therapy. The mechanistic basis of the unique associations between the abundance of IL-27 and IFNα2 and survival in only the female patients in our cohort is currently unclear and will need to be further investigated.

A few limitations should be considered when inferring the findings of our study. First, the relatively small sample size currently limits the generalizability our findings to a larger population. Further, other factors, such as ethnicity, race, type of lymphoma, type of CAR T-cell therapy and the use of bridging therapy may influence the outcome of CAR T-cell therapy response in a sexually dimorphic manner. We were unable to investigate these factors, either due to lack of available data or small cohort size. Despite these limitations, our pilot study suggests that the observed differences in clinical survival outcome in male and female patients are primarily driven by different biological interactions of cytokines and tumor microenvironment in each sex. Further research is needed to validate this hypothesis and explore the underlying mechanisms.

In summary, our study reveals sex bias in the association of pre-treatment clinical and molecular biomarkers with clinical outcomes of CAR T-cell therapy in r/r B-NHL patients, and underscores the importance of acknowledging sexual dimorphism in the risk factors associated with outcomes of personalized cell-based immunotherapies. Stratifying the outcomes separately in male and female patients may provide avenues for gender specific adaptations of CAR-T cell treatment.

## Data Availability

All data produced in the present work are contained in the manuscript

## Acknowledgments

This study was funded by VeloSano Bike to Cure, Cleveland Clinic Center of Excellence in Lymphoid Malignancies Research, and Taussig Cancer Institute. The authors thank the patients who consented to this study, as well as clinical nurses and coordinators who contributed meaningful effort to this study.

## Authorship Contributions

M.S.P. and A.J. collected and processed the blood samples; M.S.P. and A.M. collected clinical data; M.S.P., P.B., S. P., B.T.H., and N.G. analyzed the data; M.S.P., S.P., B.T.H., and N.G. wrote the manuscript; B.T.H. and N.G. provided funding support; and N.G. conceptualized and supervised the study.

## Data sharing statement

For original data, please contact the corresponding author.

## References

1. Nastoupil LJ, Jain MD, Feng L, et al. Standard-of-Care Axicabtagene Ciloleucel for Relapsed or Refractory Large B-Cell Lymphoma: Results From the US Lymphoma CAR T Consortium. J Clin Oncol. 2020;38(27):3119–3128.

2. Neelapu SS, Locke FL, Bartlett NL, et al. Axicabtagene Ciloleucel CAR T-Cell Therapy in Refractory Large B-Cell Lymphoma. N Engl J Med. 2017;377(26):2531–2544.

3. Schuster SJ, Bishop MR, Tam CS, et al. Tisagenlecleucel in Adult Relapsed or Refractory Diffuse Large B-Cell Lymphoma. N Engl J Med. 2019;380(1):45–56.

4. Crump M, Neelapu SS, Farooq U, et al. Outcomes in refractory diffuse large B-cell lymphoma: results from the international SCHOLAR-1 study. Blood. 2017;130(16):1800–1808.

5. Locke FL, Siddiqi T, Jacobson CA, et al. Real-world and clinical trial outcomes in large B-cell lymphoma with axicabtagene ciloleucel across race and ethnicity. Blood. 2024;143(26):2722–2734.

6. Mersha TB, Abebe T. Self-reported race/ethnicity in the age of genomic research: its potential impact on understanding health disparities. Hum Genomics. 2015;9(1):1.

7. Turner BE, Steinberg JR, Weeks BT, Rodriguez F, Cullen MR. Race/ethnicity reporting and representation in US clinical trials: A cohort study. The Lancet Regional Health – Americas. 2022;11.

8. Klein SL. The effects of hormones on sex differences in infection: from genes to behavior. Neurosci Biobehav Rev. 2000;24(6):627–638.

9. Afshan G, Afzal N, Qureshi S. CD4+CD25(hi) regulatory T cells in healthy males and females mediate gender difference in the prevalence of autoimmune diseases. Clin Lab. 2012;58(5-6):567–571.

10. Billi AC, Kahlenberg JM, Gudjonsson JE. Sex bias in autoimmunity. Curr Opin Rheumatol. 2019;31(1):53–61.

11. Rubtsova K, Marrack P, Rubtsov AV. Sexual dimorphism in autoimmunity. J Clin Invest. 2015;125(6):2187–2193.

12. Ribas A, Wolchok JD. Cancer immunotherapy using checkpoint blockade. Science. 2018;359(6382):1350–1355.

13. Wang PF, Song HF, Zhang Q, Yan CX. Pan-cancer immunogenomic analyses reveal sex disparity in the efficacy of cancer immunotherapy. Eur J Cancer. 2020;126:136–138.

14. Saad M, Tan AC, Naqa IE, et al. 88 Evidence of enhanced immune activation within the tumor microenvironment and the circulation of female patients with high-risk melanoma compared to males. J Immunother Cancer. 2021;9(Suppl 2):A96–A96.

15. Lefèvre N, Corazza F, Valsamis J, et al. The Number of X Chromosomes Influences Inflammatory Cytokine Production Following Toll-Like Receptor Stimulation. Front Immunol. 2019;10:1052.

16. Berghella AM, Contasta I, Lattanzio R, et al. The Role of Gender-specific Cytokine Pathways as Drug Targets and Gender-specific Biomarkers in Personalized Cancer Therapy. Curr Drug Targets. 2017;18(4):485–495.

17. Santoni M, Rizzo A, Mollica V, et al. The impact of gender on The efficacy of immune checkpoint inhibitors in cancer patients: The MOUSEION-01 study. Crit Rev Oncol Hematol. 2022;170:103596.

18. Grassadonia A, Sperduti I, Vici P, et al. Effect of Gender on the Outcome of Patients Receiving Immune Checkpoint Inhibitors for Advanced Cancer: A Systematic Review and Meta-Analysis of Phase III Randomized Clinical Trials. J Clin Med. 2018;7(12):542.

19. Burkhardt B, Zimmermann M, Oschlies I, et al. The impact of age and gender on biology, clinical features and treatment outcome of non-Hodgkin lymphoma in childhood and adolescence. Br J Haematol. 2005;131(1):39–49.

20. Morton LM, Wang SS, Devesa SS, Hartge P, Weisenburger DD, Linet MS. Lymphoma incidence patterns by WHO subtype in the United States, 1992-2001. Blood. 2006;107(1):265–276.

21. Jalota A, Hershberger CE, Patel MS, et al. Host metabolome predicts the severity and onset of acute toxicities induced by CAR T-cell therapy. Blood Advances. 2023;7(17):4690–4700.

22. Cheson BD, Fisher RI, Barrington SF, et al. Recommendations for initial evaluation, staging, and response assessment of Hodgkin and non-Hodgkin lymphoma: the Lugano classification. J Clin Oncol. 2014;32(27):3059–3068.

23. Suguro M, Kanda Y, Yamamoto R, et al. High serum lactate dehydrogenase level predicts short survival after vincristine-doxorubicin-dexamethasone (VAD) salvage for refractory multiple myeloma. Am J Hematol. 2000;65(2):132–135.

24. Karschnia P, Jordan JT, Forst DA, et al. Clinical presentation, management, and biomarkers of neurotoxicity after adoptive immunotherapy with CAR T cells. Blood. 2019;133(20):2212–2221.

25. Faruqi AJ, Ligon JA, Borgman P, et al. The impact of race, ethnicity, and obesity on CAR T-cell therapy outcomes. Blood Adv. 2022;6(23):6040–6050.

26. Liu Y, Jie X, Nian L, et al. A combination of pre-infusion serum ferritin, CRP and IL-6 predicts outcome in relapsed/refractory multiple myeloma patients treated with CAR-T cells. Frontiers in Immunology. 2023;14.

27. Vercellino L, Di Blasi R, Kanoun S, et al. Predictive factors of early progression after CAR T-cell therapy in relapsed/refractory diffuse large B-cell lymphoma. Blood Adv. 2020;4(22):5607–5615.

28. Troppan KT, Schlick K, Deutsch A, et al. C-reactive protein level is a prognostic indicator for survival and improves the predictive ability of the R-IPI score in diffuse large B-cell lymphoma patients. British Journal of Cancer. 2014;111(1):55–60.

29. Faramand R, Jain M, Staedtke V, et al. Tumor Microenvironment Composition and Severe Cytokine Release Syndrome (CRS) Influence Toxicity in Patients with Large B-Cell Lymphoma Treated with Axicabtagene Ciloleucel. Clin Cancer Res. 2020;26(18):4823–4831.

30. Jain MD, Zhao H, Wang X, et al. Tumor interferon signaling and suppressive myeloid cells are associated with CAR T-cell failure in large B-cell lymphoma. Blood. 2021;137(19):2621–2633.

31. He Y, Su Y, Zeng J, et al. Cancer-specific survival after diagnosis in men versus women: A pan-cancer analysis. MedComm (2020). 2022;3(3):e145.

32. Riihijärvi S, Taskinen M, Jerkeman M, Leppä S. Male gender is an adverse prognostic factor in B-cell lymphoma patients treated with immunochemotherapy. Eur J Haematol. 2011;86(2):124–128.

33. Bayik D, Zhou Y, Park C, et al. Myeloid-Derived Suppressor Cell Subsets Drive Glioblastoma Growth in a Sex-Specific Manner. Cancer Discov. 2020;10(8):1210–1225.

34. Kiaee F, Jamaati H, Shahi H, et al. Immunophenotype and function of circulating myeloid derived suppressor cells in COVID-19 patients. Scientific Reports. 2022;12(1):22570.

35. Jimenez RV, Kuznetsova V, Connelly AN, Hel Z, Szalai AJ. C-Reactive Protein Promotes the Expansion of Myeloid Derived Cells With Suppressor Functions. Front Immunol. 2019;10:2183.

36. Umansky V, Blattner C, Gebhardt C, Utikal J. The Role of Myeloid-Derived Suppressor Cells (MDSC) in Cancer Progression. Vaccines (Basel). 2016;4(4):36.

37. Schlecker E, Stojanovic A, Eisen C, et al. Tumor-infiltrating monocytic myeloid-derived suppressor cells mediate CCR5-dependent recruitment of regulatory T cells favoring tumor growth. J Immunol. 2012;189(12):5602–5611.

38. Salomon BL, Leclerc M, Tosello J, Ronin E, Piaggio E, Cohen JL. Tumor Necrosis Factor α and Regulatory T Cells in Oncoimmunology. Front Immunol. 2018;9:444.

39. Sanmamed MF, Carranza-Rua O, Alfaro C, et al. Serum interleukin-8 reflects tumor burden and treatment response across malignancies of multiple tissue origins. Clin Cancer Res. 2014;20(22):5697–5707.

40. Gazzaniga S, Bravo AI, Guglielmotti A, et al. Targeting Tumor-Associated Macrophages and Inhibition of MCP-1 Reduce Angiogenesis and Tumor Growth in a Human Melanoma Xenograft. Journal of Investigative Dermatology. 2007;127(8):2031–2041.

41. Mazur G, Wróbel T, Butrym A, Kapelko-Słowik K, Poreba R, Kuliczkowski K. Increased monocyte chemoattractant protein 1 (MCP-1/CCL-2) serum level in acute myeloid leukemia. Neoplasma. 2007;54(4):285–289.

42. Zhang L, Zhang M, Wang L, et al. Identification of CCL4 as an Immune-Related Prognostic Biomarker Associated With Tumor Proliferation and the Tumor Microenvironment in Clear Cell Renal Cell Carcinoma. Front Oncol. 2021;11:694664.

43. Hashwah H, Bertram K, Stirm K, et al. The IL-6 signaling complex is a critical driver, negative prognostic factor, and therapeutic target in diffuse large B-cell lymphoma. EMBO Molecular Medicine. 2019;11(10):e10576.

44. Mozas P, Rivas-Delgado A, Rivero A, et al. High serum levels of IL-2R, IL-6, and TNF-α are associated with higher tumor burden and poorer outcome of follicular lymphoma patients in the rituximab era. Leukemia Research. 2020;94:106371.

45. Koukourakis MI, Kambouromiti G, Pitsiava D, Tsousou P, Tsiarkatsi M, Kartalis G. Serum C-reactive Protein (CRP) Levels in Cancer Patients are Linked with Tumor Burden and are Reduced by Anti-hypertensive Medication. Inflammation. 2009;32(3):169–175.

46. Choi S, You S, Kim D, et al. Transcription factor NFAT5 promotes macrophage survival in rheumatoid arthritis. J Clin Invest. 2017;127(3):954–969.

47. Van Coillie E, Van Damme J, Opdenakker G. The MCP/eotaxin subfamily of CC chemokines. Cytokine Growth Factor Rev. 1999;10(1):61–86.

48. Yoshimura T. The chemokine MCP-1 (CCL2) in the host interaction with cancer: a foe or ally? Cell Mol Immunol. 2018;15(4):335–345.

49. Mittal P, Wang L, Akimova T, et al. The CCR2/MCP-1 Chemokine Pathway and Lung Adenocarcinoma. Cancers (Basel). 2020;12(12):3723.

50. Wang R, Huang K. CCL11 increases the proportion of CD4+CD25+Foxp3+ Treg cells and the production of IL[2 and TGF[β by CD4+ T cells via the STAT5 signaling pathway. Mol Med Rep. 2020;21(6):2522–2532.

51. Yang J, Hawkins OE, Barham W, et al. Myeloid IKKβ Promotes Antitumor Immunity by Modulating CCL11 and the Innate Immune Response. Cancer Research. 2014;74(24):7274–7284.

52. Conforti F, Pala L, Goldhirsch A. Different effectiveness of anticancer immunotherapy in men and women relies on sex-dimorphism of the immune system. Oncotarget. 2018;9(58):31167–31168.

53. Kim SY, Lee S, Lee E, et al. Sex-biased differences in the correlation between epithelial-to-mesenchymal transition-associated genes in cancer cell lines. Oncol Lett. 2019;18(6):6852–6868.

54. Zhu Y, Shao X, Wang X, Liu L, Liang H. Sex disparities in cancer. Cancer Lett. 2019;466:35–38.

55. Pinheiro I, Dejager L, Libert C. X-chromosome-located microRNAs in immunity: might they explain male/female differences? The X chromosome-genomic context may affect X-located miRNAs and downstream signaling, thereby contributing to the enhanced immune response of females. Bioessays. 2011;33(11):791–802.

56. Klein SL, Flanagan KL. Sex differences in immune responses. Nature Reviews Immunology. 2016;16(10):626–638.

57. Afsahi A, Burchett R, Baker CL, Moore AE, Bramson JL. Constitutive expression of interleukin-27 diminishes proinflammatory cytokine production without impairing effector function of engineered T cells. Cytotherapy. 2023;25(9):913–919.

58. Postal M, Vivaldo JF, Fernandez-Ruiz R, Paredes JL, Appenzeller S, Niewold TB. Type I interferon in the pathogenesis of systemic lupus erythematosus. Curr Opin Immunol. 2020;67:87–94.

