## Supplemental Materials for "Sex-specific Risk Factors for Survival in B-cell Non-Hodgkin Lymphoma Patients after Anti-CD19 CAR T-Cell Therapy"

**Running Title:** Sex-specific risk factors in CAR T-cell therapy

**Keywords:** CAR T-cells, survival, sex, cytokines, tumor burden

### Supplemental Figure 1

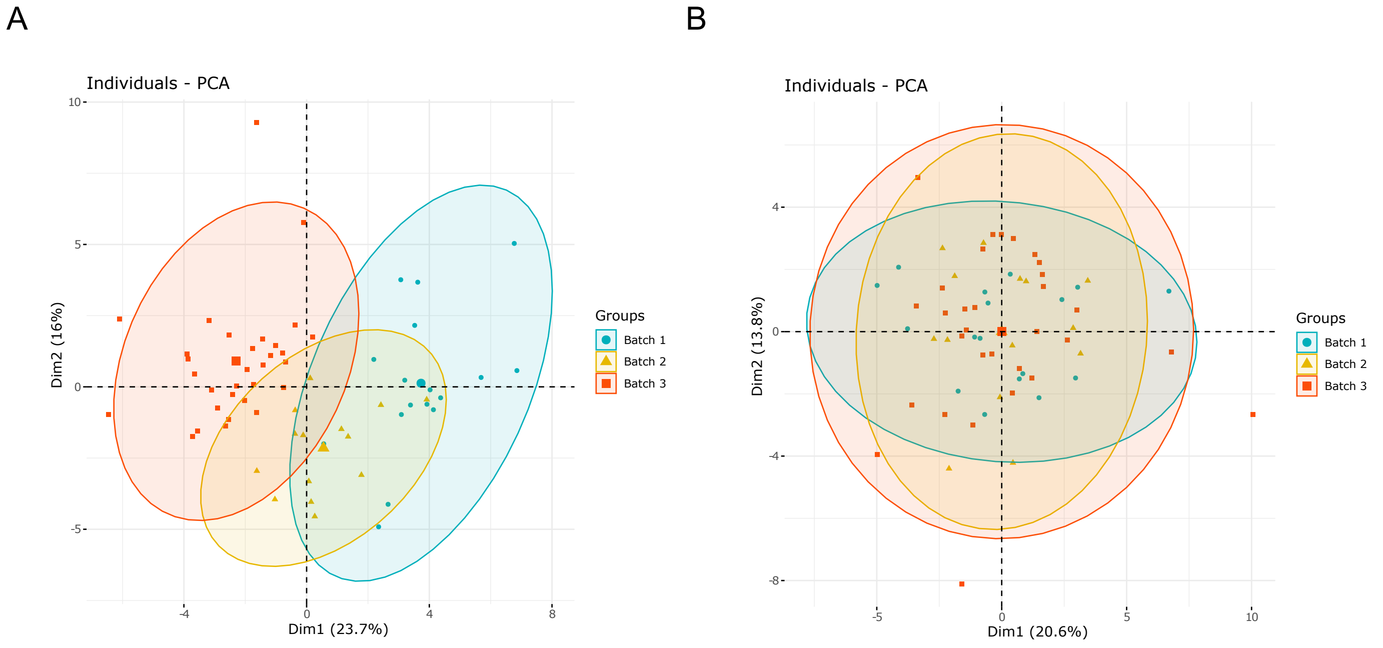


**Supplemental Figure 1. Principal component analysis of all samples quantified in three different batches. (A)** Before batch correction, **(B)** After batch correction.

#### Supplemental Figure 2

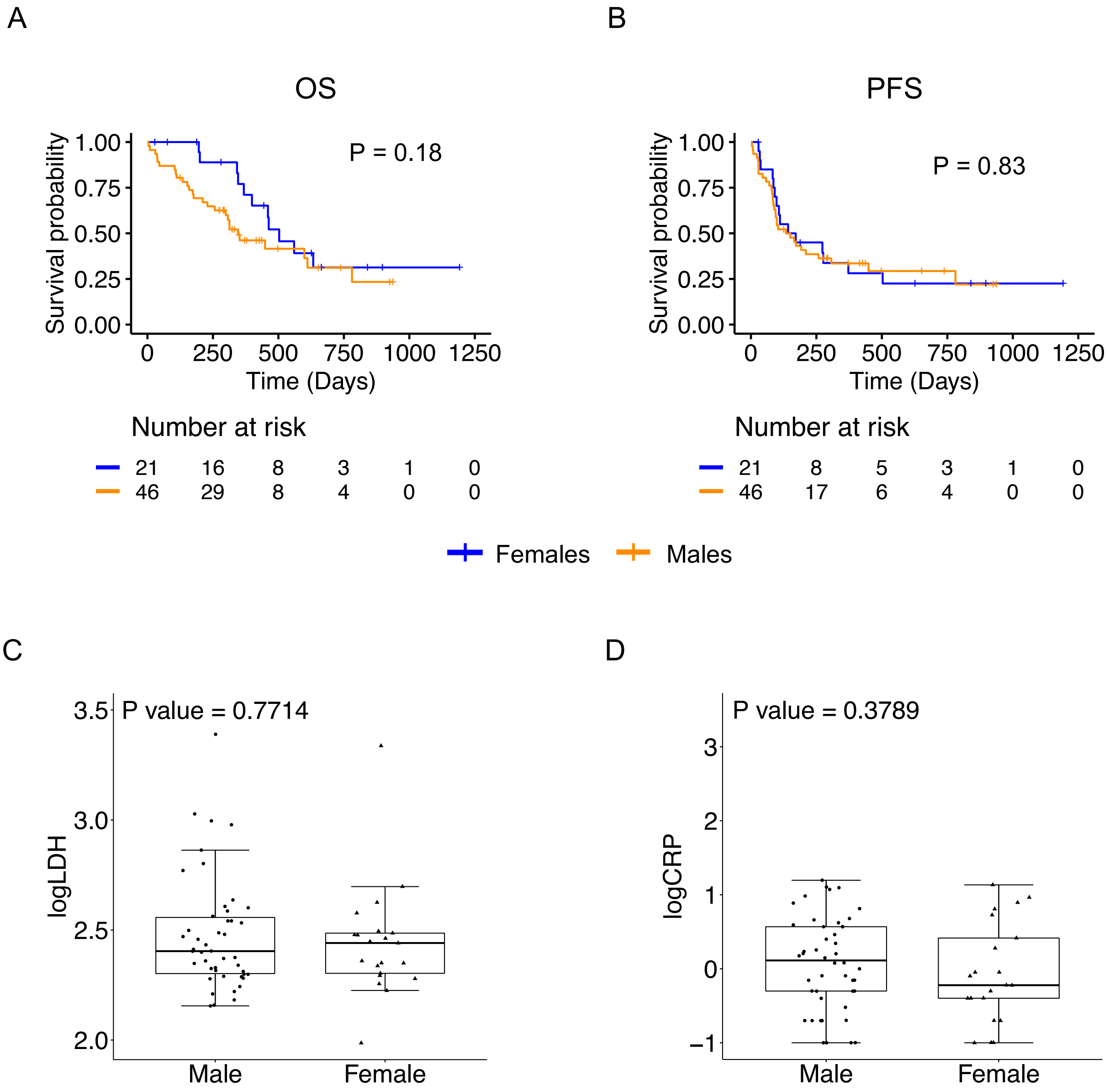


**Supplemental Figure 2. Survival, tumor burden and CRP in male and female LBCL patients.** Kaplan Meier analysis of OS **(A)**, and PFS **(B)**, and pre-treatment relative abundance of LDH **(C)**, and CRP **(D)** in male and female patients. Males, n=46; Females, n=21. Yes, n=45; Kym, n=22.

#### Supplemental Figure 3

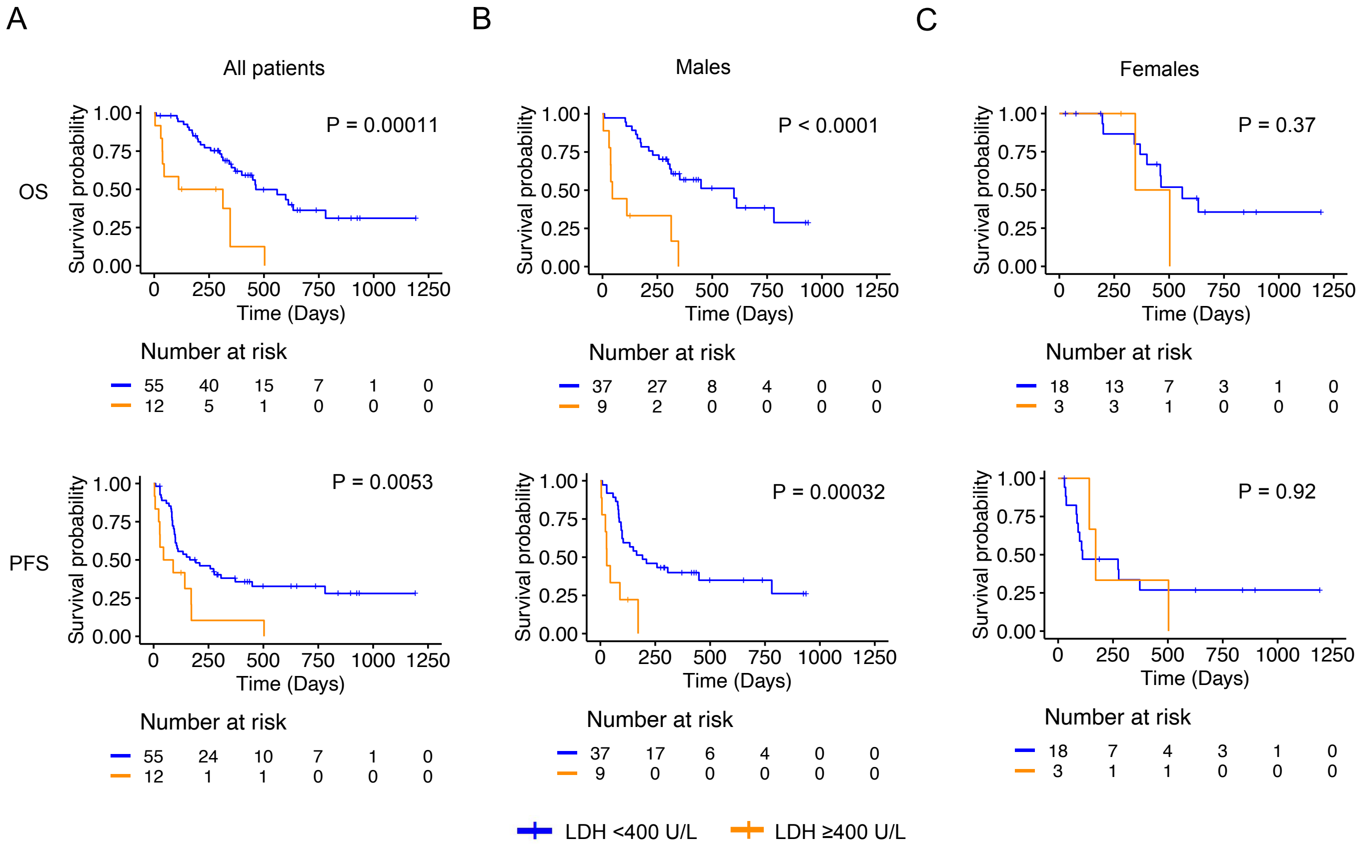


**Supplemental Figure 3. Male patients with higher pre-treatment tumor burden show poorer survival after treated with anti-CD19 CAR T-cells.** Association of baseline LDH levels with overall survival (OS), and progression-free survival (PFS) in both male and female **(A)**, only male **(B)**, or only female **(C)**, LBCL patients. P<0.05; Males, n=46; Females, n=21; Yes, n=45; Kym, n=22. LDH cut-off, 400 U/l.

### Supplemental Figure 4

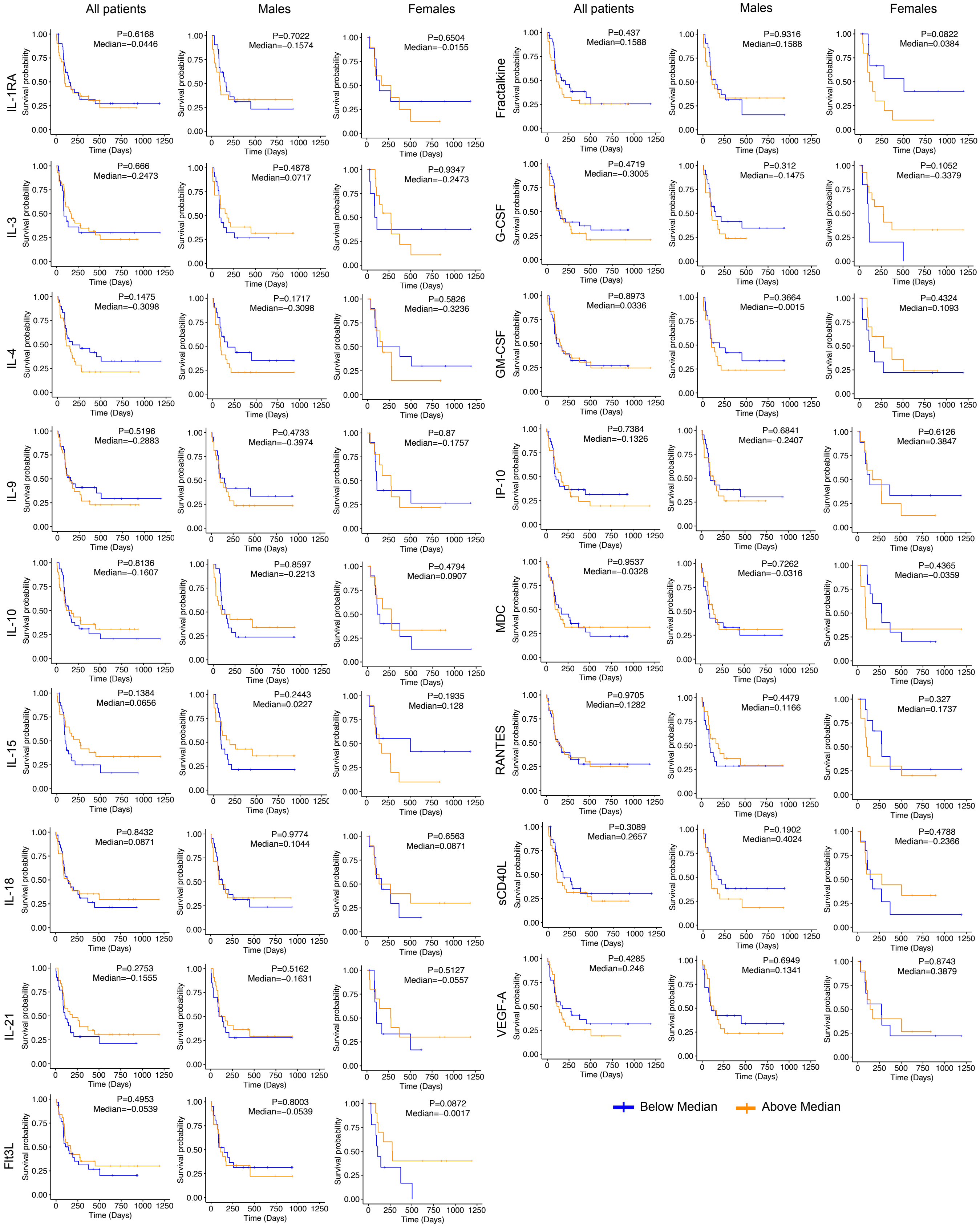


**Supplemental Figure 4. Lack of association between the relative abundance of a subset of cytokines/ chemokines and PFS in LBCL patients receiving anti-CD19 CAR T-cell therapy.** Association between relative abundance of the indicated cytokines and PFS in all, male or female LBCL patients. ($P<0.05$), Males, n=42; Females, n=20; Yes, n=42; Kym, n=20.

### Supplemental Figure 5

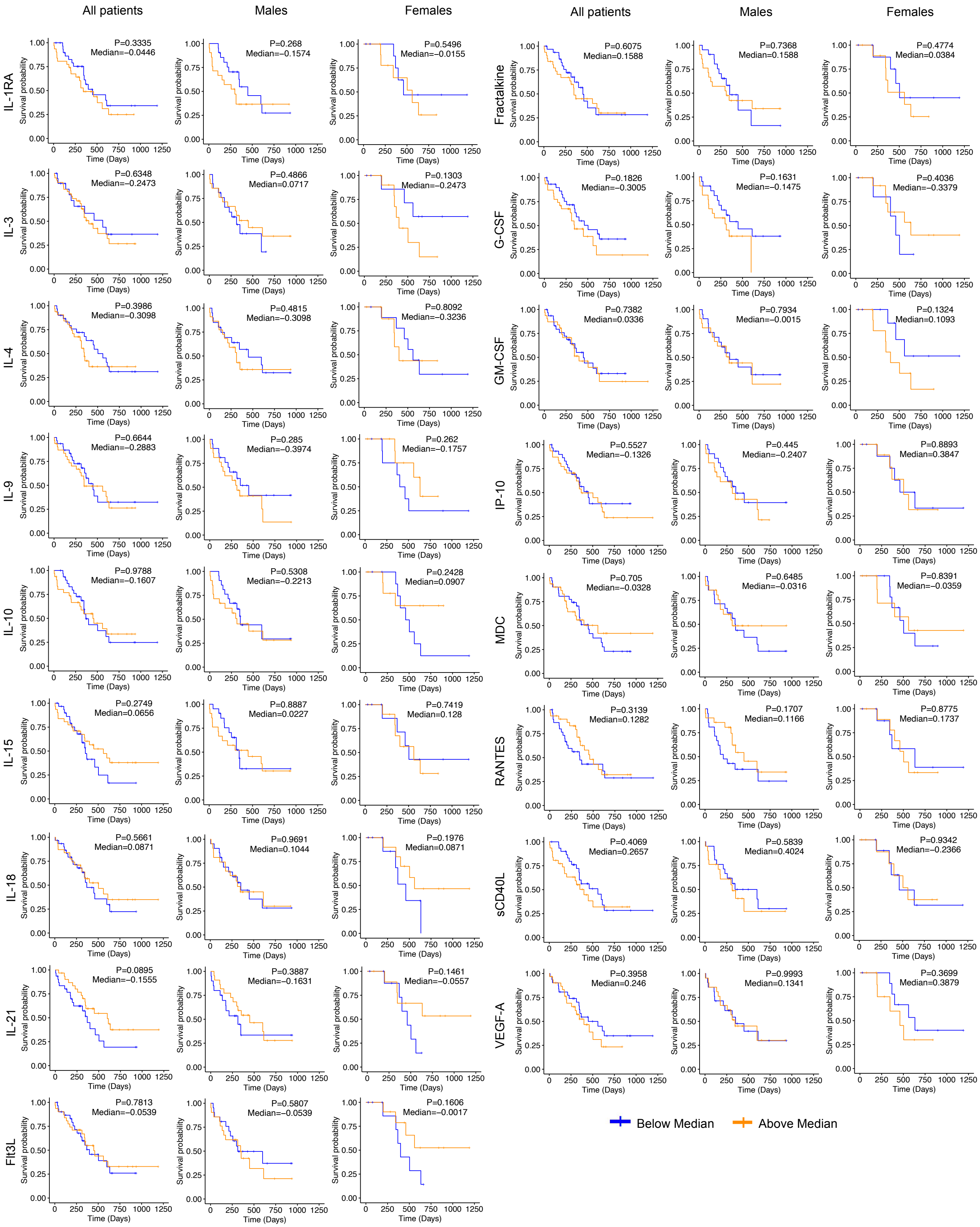


**Supplemental Figure 5. Lack of association between the relative abundance of a subset of cytokines/ chemokines and OS in LBCL patients receiving anti-CD19 CAR T-cell therapy.** Association between relative abundance of the indicated cytokines and OS in all, male or female LBCL patients. (P<0.05), Males, n=42; Females, n=20; Yes, n=42; Kym, n=20.
